# *Instead of seeing it as a health care issue, you see it as you:* Reasons for alcohol use, consequences of use, and barriers to help seeking among fathers in Kenya

**DOI:** 10.1101/2024.06.25.24309498

**Authors:** Ali Giusto, Emily N. Satinsky, Florence Jaguga, Wilter Rono, Julius Barasa, Chardée A. Galán, Milton L. Wainberg

## Abstract

**Introduction:** Father alcohol use negatively impacts family systems, yet research in this area is scarce in low- and middle-income countries like Kenya. An understanding of why fathers drink, consequences of alcohol use, and barriers to care is needed to refine and adapt clinical and implementation approaches to treating fathers.

**Methods:** Community members, leaders, mental health providers, and fathers experiencing alcohol use problems in Eldoret, Kenya were recruited to participate in semi-structured qualitative interviews and focus groups. Participants were asked about why fathers engage in alcohol use, potential impacts of use, and barriers to accessing care. The frame method was employed to analyze the data. The study team read transcripts, iteratively memo-ed and discussed notes, developed a codebook, and coded transcripts. Broad codes were summarized and reviewed alongside transcripts.

**Results:** Participants noted reasons for and consequences of fathers’ drinking at individual, family and interpersonal, and sociocultural levels. At the individual level, alcohol use facilitated an escape from mental distress and acted as a means to cope with “idleness” due to unemployment. Consequences included poor physical and mental health, such as depression. At the family/interpersonal level, fathers used alcohol to distract themselves from family conflicts. Consequences included violence and poor child outcomes. Gender and drinking norms were drivers at the sociocultural level. Consequences at this level included stigmatization and loss of social status, which can drive shame and isolation. Salient barriers to care included fathers’ lack of awareness of their alcohol use problem, limited-service access, and social stigma.

**Conclusions:** Father motivations for drinking are influenced by multiple ecological levels, and drinking has a cascade of consequences on the family. These effects are worsened by barriers to care. Intervention and implementation strategies should consider masculinity norms, resources, and avoidant coping motivations in adaptation.

## Introduction

Globally, alcohol use disproportionally impacts men. Individual and family consequences of alcohol use account for 9.6% of disability adjusted life years (Rehm et al., 2009, 2010). In Kenya, the prevalence of alcohol use disorders (AUDs) is higher in men than women (15.7% men, 4.6 % women) (NACADA, 2022). Further, in Kenya, the population experiences the third highest total disability-adjusted life years from AUD in Africa (Degenhardt et al., 2018). Alcohol use is often comorbid with depression, bipolar disorder, and anxiety, such that problems with alcohol are sometimes seen as a more diffuse marker of mental health problems (Jane-Llopis & Matytsina, 2006; Kendler et al., 2003). This is particularly common among men who tend to underreport internalizing mental health issues like depression (Sigmon et al., 2005). Furthermore, men’s alcohol use is associated with increased risk of mortality, cancer, poor mental health, and violence (Carvalho et al., 2019; Roerecke & Rehm, J., 2013).

The consequences of men’s alcohol use extend beyond the individual to impact their families (Flouri & Buchanan, 2003; Sigmon et al., 2005). Alcohol use among men, and specifically fathers, can disrupt family systems and impart negative impacts on the couple relationship, the parent-child relationship, and family member mental health (Goeke-Morey & Mark Cummings, 2007; Rossow et al., 2015). Although research has explored factors associated with father alcohol use and its consequences, this research has primarily been conducted in high-income, Western settings like the United States. In low-and middle-income countries like Kenya, these issues remain understudied (Jaguga & Kwobah, 2020a). More research into the consequences of men’s alcohol use in Kenya is needed given disproportionate alcohol use among men compared to women as well as the important role fathers play in families in this setting (Giusto et al., 2021; Jaguga & Kwobah, 2020; NACADA, 2022). Furthermore, fathers are central to family and family member well-being, but they are often missing from the family-focused literature (Flouri, 2010; Giusto et al., 2017). This is true in Kenya and globally (Panter-Brick et al., 2014). As such, there is a clear need to explore fathers’ experiences and engage them in care alongside female caregivers, where much of the research is currently focused.

Understanding fathers’ motivations for engaging in alcohol use, as well as its impact on their mental health and family dynamics, is crucial for clinical assessment, treatment, and the development of targeted and contextualized interventions. Most non-pharmacological treatments for alcohol use include psychoeducation and assessment of drinking motivations to replace those behaviors (Yeh et al., 2017). Qualitative methods present an opportunity to explore how community members view reasons for and consequences of fathers’ alcohol use. This thereby allows for a rich, nuanced picture of fathers’ experiences and informs the development and adaptation of clinical approaches. For example, some qualitative evidence from Kenya suggests fathers experiencing alcohol use problems may sometimes accept informal help from community or family members but rarely seek help from formal care systems like public health facilities (Jaguga & Kwobah, 2020b; Patel et al., 2020). Continuing to specify and identify barriers that hinder fathers from seeking help or engaging in care can inform the development of strategies aimed at mitigating these problems.

The objective of this paper is to explore the reasons behind fathers’ alcohol use in Eldoret, Kenya; the consequences of fathers’ alcohol use; and the barriers that impede fathers’ help-seeking and maintain patterns and consequences of alcohol use. We explore these aims from the perception of multiple stakeholders including fathers experiencing problem alcohol use, hospital leaders, policy makers, mental health providers, and others. The broader goal of this paper is to inform clinical and implementation strategies in the area to better engage and treat fathers’ alcohol use in ways that are relevant and align with existing community strengths.

## Methods

We conducted a qualitative study with community members, leaders, mental health providers, and individuals experiencing alcohol use problems in Eldoret, Kenya. Semi-structured interview guides included questions on participants’ perceptions of alcohol use among fathers, consequences of alcohol use, current services, and barriers to help-seeking. Questions were asked as part of a larger study (Giusto et al. 2023) focused on identifying implementation determinants for delivering a mental health and alcohol use intervention for fathers in the area. This study was guided by the Consolidated Framework for Implementation Research (CFIR) and Integrated Sustainability Framework (ISF; Damschroder et al., 2009; Shelton et al., 2018), which helps organize questions by various domains often implicated in implementation such as the outer-level (i.e., culture, stigma, country), inner-level (i.e., settings like hospital or community), individual-level (i.e., characteristics, attitudes believes or providers and patients). The study is reported following the Standards for Reporting Qualitative Research (Appendix Table 1;).

### Location

Interviews and focus groups were conducted in Eldoret, Kenya, in partnership with the Kenyan Ministry of Health, Moi Teaching and Referral Hospital (MTRH), and AMPATH, a collaborative service and research entity comprising MTRH and a consortium of North American educational institution (https://www.ampathkenya.org/). Ethical approval was obtained from the Institutional Review Board at New York State Psychiatric Institute (Protocol 8084, Date Approved 29 October 2020) and the Institutional Ethics Review Committee at Moi Teaching and Referral Hospital (Approval Number 0001138, Date Approved: 10 December 2020). Verbal and written consent was obtained from all participants.

### Participants

Participants included individuals with a range of experiences with alcohol use and mental health treatment in the area. A total of 18 individual key informant interviews (KIIs) and eight focus group discussions (FGDs; N: 31 participants) were conducted. KII participants included four community leaders, three policy makers, four hospital leaders, five mental health and alcohol use providers, three individuals who received alcohol use treatment, and three peer-father counselors (i.e., lay counselors trained to provide alcohol use treatment). FGDs were conducted with fathers experiencing alcohol problems (6 participants, 1 FGD), mental health providers (21 participants, 3 separate FGDs), and community leaders (4 participants, 1 FGD). A diverse selection of participants was sought, including individuals from various sectors, both rural and urban, and lay and professional backgrounds. Individuals currently experiencing alcohol use problems were eligible if they were fathers responsible for children under 18, exhibited alcohol use problems (indicated by Alcohol Use Disorder Identification Test, score of 8-19 (Babor et al., 2001)), and had engaged in alcohol use within the past two months. Leaders, providers, previous peer-father counselors, and fathers who previously received alcohol use treatment were identified either through study team knowledge and/or previous study records (if individuals indicated willingness for future contact). All participants were included if they were interested and consented to participate (a full description of procedures can be found in [blinded]).

### Recruitment

Potential participants were approached and consented by a trained Kenyan research assistant (RA) or the Project Coordinator. Recruitment occurred from March 1, 2020 to March 15, 2022 (please note there were delays due to COVID). Community leaders and providers were identified based on study team knowledge and consultation with local leaders; they were contacted by phone, email, or in person to assess initial interest. If the community leader or provider expressed interest, the RA or project coordinator obtained consent.

Previous patients and peer-father counselors were identified through records of a previous project led by the lead author. Those who expressed interested in being contacted for future studies were then randomly assigned identification numbers. They were approached in order of their ID number and invited to participate in this study.

Current patients were recruited when presenting to [name blinded] or affiliated support groups. They were approached by an RA who explained the study. If individuals expressed interest, the RA completed the screening process and consented eligible fathers. Fathers who did not meet eligibility criteria were assured that they would continue receiving care with [name blinded].

### Procedures

This study took place in 2021 to 2022. Data collection was conducted by Kenyan RAs in collaboration with the on-site investigator. The on-site investigator had a relationship with some of the policy makers and mental health providers. In these cases, the interview was conducted by the team member who knew the participant least. Interviews and FGDs were conducted in English or Kiswahili based on participants’ preferences. Key informant interviews typically lasted around 45 minutes. Focus group discussions were longer and typically took between two and three hours. All sessions were recorded, deidentified, and transcribed into English. Specific Kiswahili idioms were maintained in the transcripts as appropriate. (The full interview guides are available in the larger study paper, [blinded]).

### Analysis

Qualitative analysis was guided by the framework method (Gale et al., 2013). The framework method groups data into codes that are used to manage and organize the data. It generally has seven stages starting with transcription and ending with interpretation. We incorporated both data-driven and predefined codes based on implementation framework domains as well as the questions asked (e.g., reasons for engaging in alcohol). The analysis team included a Kenyan psychiatrist, a Kenyan research manager, and a US-based psychologist. This allowed us to interpret and balance data from multiple, diverse perspectives.

The team read the transcripts and took notes in memos. The team then discussed notes to generate broad codes. Next, individual team members coded a subset of randomly selected interviews by interview type. Use of codes and transcripts were discussed to develop a coding framework and codebook. Transcripts were coded independently until 80% agreement was met across four coders. Since percent agreement is directly interpretable (McHugh, 2012), we chose this method to determine when to move to independent coding. Next, transcripts were divided and coded independently using NVivo 12.0; questions that arose were discussed and consensus reached. Coded data were then reviewed and summarized. Lastly, we interpreted data. As these aims focused on the reasons and consequences of alcohol use as well as barriers that might influence these factors, data were organized by these codes. Finally, codes were synthesized with reference to transcripts as needed.

## Results

Across participants, multiple interrelated factors were described as driving father’s alcohol use and the consequences of use. Although alcohol use treatment services exist in the region, participants described barriers that stopped fathers from seeking and accessing care. These barriers appeared to maintain and reinforce patterns and consequences of use. Figure 1 provides an overview of these results. Below, we first describe noted reasons for drinking, and then we describe consequences of fathers’ alcohol use. These themes occurred within different ecological levels that differed somewhat from the domains explicated by the implementation frameworks. The levels that emerged from the data were at the individual level, family and interpersonal level, and sociocultural level. We then present barriers to treatment that specifically impacted fathers’ reasons for drinking and maintained consequences of use.

**Figure 1.**
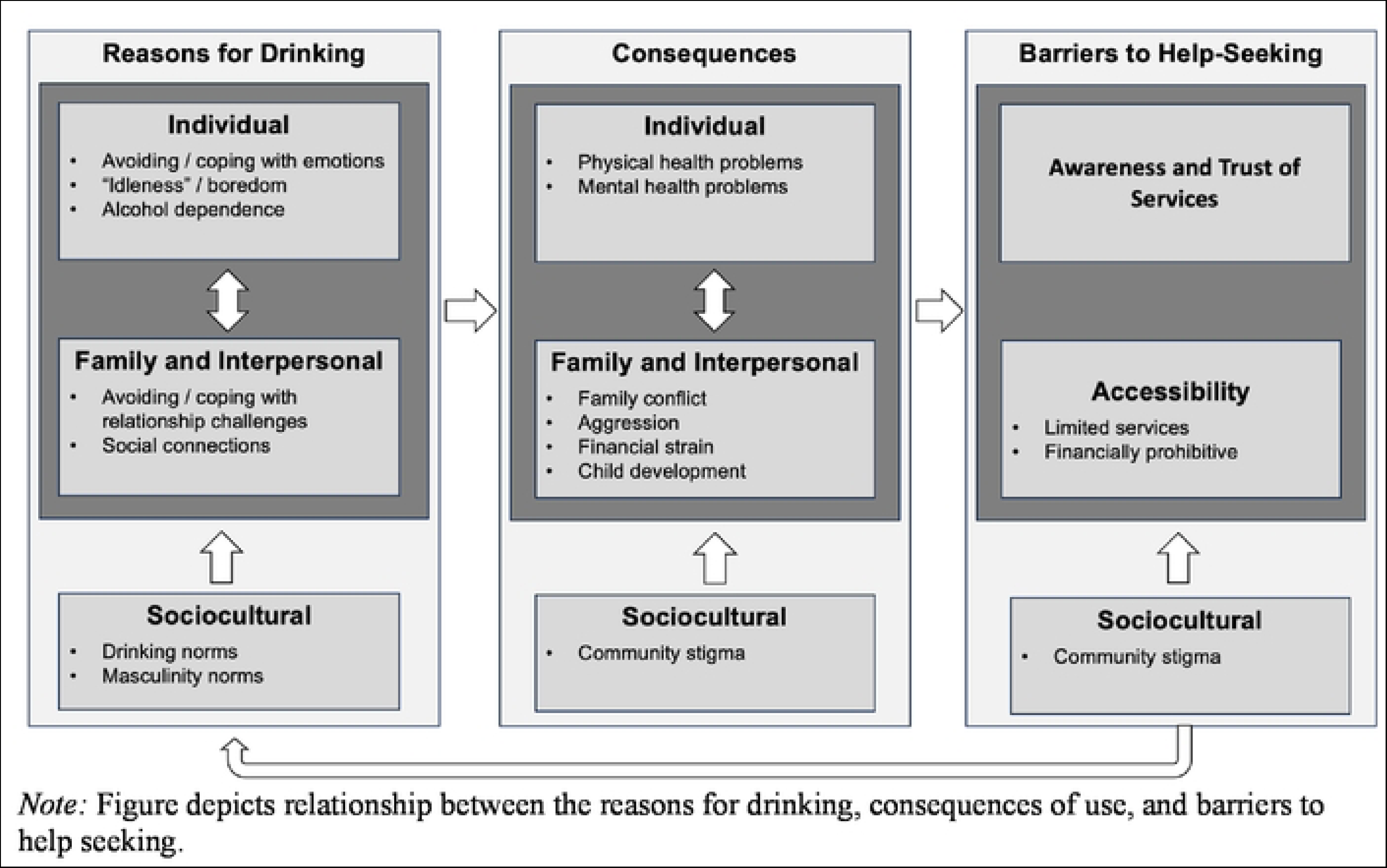
Interrelated Cycles of Reasons for Drinking, Consequences, and Barriers to Help Seeking among Fathers in Kenya

### Reasons for drinking

Reasons for fathers’ drinking were noted at multiple levels. The function of drinking— i.e., to avoid emotion or connect—was often similar across these levels. At the individual level, stressors included mental health problems, “idleness”, and dependence. Interpersonal factors included family and peer conflict as well as social connection. Sociocultural drivers included norms around drinking and cultural expectations regarding masculinity. We organize results by these levels.

#### Individual

Participants often cited mental health problems or difficult emotions as key drivers of alcohol use. Specific concerns included depression, anxiety, stress, trauma, anger, grief, or poor coping skills and the use of alcohol as a means to escape these problems (i.e., avoidant coping). Many participants noted that alcohol use to escape mental distress can be driven by financial stress occurring in a context of poverty and expectations to financially provide. One community leader described, “*you find that someone is worried. They have a lot of bills to take care of,* [and] *the relationship problems…they want to forget about all these problems, and so what do they do? The next solution is alcohol*” (FGD 101 – P1). Similarly, two fathers currently in treatment described using alcohol to cope with anger. One described,“*something small can make me angry. You run away telling yourself that you want to go and take away the anger*” (KII 1603). However, this same patient noted that alcohol does not fix the anger – “*you get angry again*.”

Related, drinking to cope with idleness or boredom emerged as a key theme. Participants described how boredom and feeling a lack of purpose, often in the context of unemployment, led fathers to drink. Some participants noted how COVID-19 amplified this problem. Many fathers lost work during the pandemic and turned to alcohol to cope with “idleness”. One participant described, “*They do not have anything to do in terms of work. Most of them are jobless and so they wake up in the morning and…they tend to take alcohol because they do not have anything to do*” (KII 1302 – Clinical Officer).

Lastly, physical dependence on alcohol, or reliance on alcohol to function, was noted as both a reason and consequence of alcohol use. Patients and providers described how fathers might start drinking as soon as they wake up to get energy to start the day.

#### Family and Interpersonal

Challenges in fathers’ family and peer relationships were also cited as reasons for drinking. Similar to dealing with negative emotions, participants described how fathers use alcohol to avoid conflict and forget quarrels with partners or difficult interactions with their children. For instance, one participant reported: “*Marital conflict is a key thing. And once you have had conflict, you do not wish to go back to the house and face the same woman you fought, and so what do you do? You go there to the* [alcohol woman] *and forget yourself for a moment*” (FGD 101 – P1). Conflicts outside the immediate family were also noted as reasons for drinking. A counseling psychologist, for example, shared, “*The other challenge is…the relationship at the workplace…and in the community generally*” (KII 1309).

While many participants described alcohol as a way to escape, some participants shared that fathers might use alcohol to handle problems directly. These participants reflected that some men feel more comfortable confronting their wives or talking through disagreements when they are drunk. A current patient described, “[I] *cannot confront the problem when I am not drunk*”; the same patient added that when he is drunk, “*Whatever was bothering me will all come out…When you drink alcohol, you say it all*” (FGD 701 – P3). Other participants discussed a similar sentiment in reference to peers—alcohol gives men the courage to confront community members who wronged them or talk openly with peers about marital stress.

Lastly, alcohol use was described by some as a means of social connection, such that a reason for drinking may be celebrating or being with friends. A psychiatrist described these phenomena saying, “*It is like a support group kind of… so they form these cocoons, and so these cocoons propel the drinking. They help each other drink. If you do not have money, I will buy for you today, if you have tomorrow, you will buy for me…so they form a cocoons of similar behavior. They live with those cocoons until when they have advanced issues*” (FGD 401 – P3).

#### Sociocultural

Outer-level factors related to gender norms and drinking norms emerged as key drivers of father alcohol use. These intersected with and compounded individual and interpersonal level drivers by reinforcing and contributing to motivations for using alcohol as a means to escape or cope with distressing emotional experiences.

##### Drinking norms

Participants described two social factors that promote fathers’ alcohol use: peer pressure and cultural events tied to alcohol. First, one participant noted, “*we take it for granted that the peer pressure affects the youth alone, but I have seen it affect even men*” (FGD 101 – P2 youth student leader). Men engage in alcohol use when they see it modeled in others, with one individual noting, “*That issue of being* [the] *odd one out can become difficult*” (FGD 702 – P3 patient). Some peer pressure may, in part, be driven by cultural norms around men’s drinking. An addiction specialist described, “*culturally, you are not supposed to say that I cannot drink*” (KII 1203). A psychiatrist added, “*in the norms of many societies, they feel that if a man does not drink, then he is the odd one out*” (KII 1313).

Second, during certain events, alcohol is widely available and encouraged among men. In some areas, such as the urban informal settlements, boys and young men sell alcohol to earn a living. They drink “*because it is just there in plenty*” (FGD 401 – P1 social worker). This participant added that the presence of alcohol in these settings means that drinking often passes down through generations. Another participant mentioned that Friday is a drinking day in the community, while several others added that alcohol is embedded into “circumcision time” and other rituals. Alcohol use norms during these ceremonies can amplify social pressures to drink.

##### Masculinity Norms

Norms around masculinity emerged as a specific reason that fathers use alcohol to cope with distressing emotions. Participants repeatedly discussed expectations that men appear strong, act “macho”, or have “alpha energy”. As a clinical psychologist described, “*There is the cultural aspect where men have himself together. Man is not supposed to say that I am overwhelmed*” (KII 1203). Expectations that men never show weakness can compound feelings of shame and loneliness, and thereby prompt engagement in alcohol use. As gender norms are deeply ingrained in society, fathers often lack role models for how to deal with stress and difficult emotions in healthy ways. Men see examples of women discussing mental health problems but do not see such modeling among other men. When individuals approach their own fathers, cultural beliefs are reinforced:

> “I think it is harder for our fathers to approach their fathers. This is because when you look at those older men, they are those kind of people who are closed off…They will tell you to ‘struggle like a man,’ and so the moment you lack a role model, you will go to your fellow man whom you know…the first thing he will do is go to the bar, and so you will follow each other to the bar” (FGD 101 – P2 student leader).

Given this cultural context, men “*just lock it in*” and can turn to alcohol use and other harmful behaviors as an outlet for their emotions.

Many participants described how these problems start with “*negligence of the boy child*”. They noted that while significant funding has gone into supporting and empowering girls, boys get left behind. A psychiatric nurse elaborated,

> “We are putting more emphasis on girls; we are protecting girls more than we are protecting boys. During the growing up of the boy, we are telling them that the boy does not cry and therefore we are telling them untrue and then we are not preparing them for the challenges that are there right now…And then because of this empowerment of the girl child, the boy has lost his identity. Now, when he becomes a man or when he becomes a father or a husband, the nature of the man, is already undermined…now they enter into… alcohol. They go in groups of men and from there they feel better, they feel accepted, when they are in a group and then they are drinking. But they do not know that that kind of drinking has its own steps from curiosity up to the time the person is hooked he cannot go back. The challenge started from when he was a boy” (FGD 401 – P4).

This quote highlights how masculine norms, social connection (interpersonal), and “acceptable” coping skills (individual) can also intersect to influence and maintain men’s alcohol use. Further, participants noted that efforts to empower girls have shifted norms. A psychiatrist and social worker both commented on how women no longer depend on men financially. These changes can instill a disconnect between perceptions of societal gender roles (i.e., men should provide) and actual behaviors (i.e., women earn more). As a psychiatrist noted, this cultural conflict can further contribute to alcohol use among fathers.

### Consequences of drinking

As with the drivers of fathers’ alcohol use, the reported consequences of drinking also spanned multiple ecological levels. Participants discussed negative impacts at the individual, interpersonal, and sociocultural levels.

#### Individual

At this level, consequences included physical and mental health problems. Physical health consequences included direct effects of alcohol use on health, such as liver cirrhosis, cancer, and death. Participants also mentioned indirect effects. For example, when fathers drink, they are less likely to maintain their hygiene, thus allowing opportunistic diseases. Furthermore, many noted drinking can increase likelihood of risk behaviors that increase susceptibility to COVID-19, HIV, and other sexually transmitted infections.

Discussing mental health, participants described how alcohol use can contribute to shame, depression, and anxiety among fathers. A consultant psychiatrist commented, “*and then anxiety related to alcohol use – predisposing and also an outcome from alcohol use*” (KII 1206). Fathers’ lives become unmanageable, and they feel powerless to cope with daily life. Some providers noted that this hopelessness can also motivate suicidal ideation and behaviors.

#### Interpersonal

Participants universally noted that fathers’ excessive alcohol consumption leads to family conflict. Several participants shared that problematic alcohol use among fathers contributes to potential for physical and emotional aggression in the family, escalating at times into violence. A church leader stated, “*before they are drunk, they are just good people…but after drinking, they become violent. They become abusive*” (KII 1103). Risk of emotional and psychological abuse was also noted. Living in this environment, “*children live in fear, women live in fear*” (FGD 102 – P1 church leader). Some women threaten to leave, leading to “*breakage of the marriage*” (KII 1104 – advisory committee member).

Financially, fathers’ alcohol use constrains the availability of resources for essential needs within the family. Participants shared that when fathers engaged in problematic alcohol use, families often struggle to cover expenses for rent, school fees, medical services, seeds and fertilizers, soap, food, and sugar. As one community provider explained, some fathers even resort to selling items from their household in exchange for money to fund their drinking. A father seeking treatment for his alcohol use shared that “[you are] *entertaining yourself as your children struggle*” (KII 1603). When families are restricted in these ways, they seek money from other sources: “*the mother will result into looking for money from another man…The children can go stealing so that they can get something to eat.”* (KII 1101 – village elder).

Alcohol use was also reported to indirectly impact financial stress through professional problems. More specifically, alcohol use may result in fathers missing work, which not only threatens a family’s capacity to fulfill their essential needs on days when fathers are absent but also heightens the risk of long-term unemployment. Losing a job then amplifies conflict and financial stress in the family. A provider noted,

> “Most employers do not understand alcoholism as a disease, and they sack you. When they sack you, you go back to the house and your wife doesn’t want to understand. [She] wants to blame you for being irresponsible and abusing…your wife abandons you and even sends you away from your family because even now, you are good for nothing husband and now you get into economic hardship” (KII 1202).

Interviews highlighted the ways in which youth are affected by their fathers’ alcohol use and the resulting family conflict that it provokes. Participants shared that children in these environments lack confidence and feel depressed. Many children do not go to school consistently due to lack of school fees and familial stress. When they *do* attend school, they often perform poorly. As a church leader described, “*even if they go to school, they cannot afford to go to school five days in a week…you find that in such areas, once a man has lost focus in the family, everything scrambles in the family and in the neighborhood*” (FGD 102 – P1). In addition to poor school attendance and performance, some children show behavioral problems. A father in treatment for alcohol use reflected, “*First it hurts the children, because these children will not have discipline because when the father is not around, sometimes a mother has to deal with grown up boys. They will not respect the mother the way they respect the father. So, the children will break the rules…they become poorly disciplined.*” (FGD 702 – P2).

#### Sociocultural

When fathers’ alcohol use advances to more severe levels, there can be consequences in the community. Fathers may be stigmatized, lose social status, and get “*left behind*”. Referencing the loss of status, a mental health provider described, “*and that loss is not easy to navigate…they rather resign to fate of being called a drunkard because they can no longer stand up to pick up their position*” (FGD 401 – P5). A psychiatrist elaborated on the process by which men experiencing problems with alcohol use are pushed to the outskirts of society: “*they are shunned right from the working areas to the community activities, leadership roles in the society…they are not valued anymore. They feel that they are appendages in the community building*” (KII 1313). In this context, fathers may feel shame and further self-isolate to avoid societal humiliation. A father receiving treatment for alcohol use stated, “*you look like a bad man, and when a man looks bad…do you know that they will throw you away?…Things become so bad*” (FGD 701 – P2). Given expectations around masculinity, there appears to be little cultural empathy for men with mental health and alcohol use problems.

### Barriers to Care and Maintenance of the Cycle

Participants listed several barriers that stand in the way of men accessing alcohol use treatment or other forms of care. These barriers maintain and reinforce drinking habits and exacerbate consequences in a cyclical manner.

#### Lack of awareness and trust

Among fathers, lacking awareness of the problem and available services were barriers to help-seeking. Two psychiatrists reflected that fathers are often unable to recognize when alcohol use interferes with their life. A third psychiatrist reiterated this, adding that the lack of awareness may be due to cultural beliefs: “*You know sometimes we have cultural explanation of challenges. If you have a challenge, instead of seeing it as a health care issue, you see it as you*” (KII 1308). If fathers think that their behaviors are normative in context, they are unlikely to seek care.

A hospital manager added that fathers might also lack knowledge of available services to support them in addressing their alcohol use problems. While many women-specific health services exist, men often do not know of similar services tailored to men and may assume services are non-existent. When fathers *do* know about services, a lack of trust emerges as a further hindrance to help-seeking. Some men feel like the services cannot help them while others feel wary sharing sensitive information. For example, a psychologist mentioned that some fathers “*do not really want to express some of the issues, especially with marriage. They do not want to talk about it*” (FGD 403 – P3).

#### Accessibility barriers

Even when fathers are aware of services, providers noted that access is limited. One provider noted that the establishment of alcohol use programs is not prioritized. Distance to existing treatment programs further impeded access. Lastly, the cost of treatment arose as the most prominent accessibility concern. Some participants mentioned that, even if treatment costs are reasonable, fathers would rather spend that money on their family. A hospital manager elaborated, “*it will be down the ladder…because the child has to go to school, let me first sort out the school fees. We are lacking food here, and then, by the time you reach to yourself, I cannot pay for that service, so I will survive like a man*” (KII 1301). A psychologist added, “*they will feel that they are leaving a gap at home…who is paying the rent for those three months while I am in rehabilitation in the inpatient care?*” (FGD 402 – P1). A couple participants also mentioned potential issues with insurance.

#### Social stigma

Stigma was identified as a critical barrier to help seeking. A psychiatrist noted that drinking is not seen as a health problem but rather as a moral issue. Others described stigma surrounding the idea that people who choose to drink bring the problem upon themselves. At the same time, policies that criminalize drug use and suicide make help-seeking particularly difficult for individuals with comorbid problems. In this context, fathers might avoid care.

Social stigma is exacerbated by gender norms, limiting help-seeking among men. Referencing cultural expectations around masculinity, several participants mentioned that men are perceived as weak if they access mental or behavioral health treatment—“*you are a less of a man according to them if you go*” (KII 1309 – counseling psychologist). A rehabilitation provider summarized,

> “We are in a patriarchal environment whereby a man is supposed to be the provider…So now when a man has mental issues or is a man struggling with alcoholism, it means that the leadership role has demeaned…The stigmatizing words you hear from colleagues, friends, and family about this person…they take away the ego of a man…How can you actually confess that you are an alcoholic?…I think stigma has been the main challenge for men to seek for services” (KII 1202).

A mental health provider echoed the idea that men are supposed to be the ones with the solutions to problems rather than the ones with the problems themselves. These ideas often become internalized as shame self-stigma, further contributing to a lack of help-seeking. As a youth student leader described, “*most of them believe that help is for the weak and no one wants to be seen as weak…you know men have an ego, and so the moment you go to someone asking for help…they will say I have failed*” (FGD 101 – P2).

## Discussion

This study took a qualitative approach to understand the motivations behind alcohol consumption among fathers in Eldoret, Kenya, as well as the consequences of their alcohol use and the barriers they face seeking help. The study found that reasons for and consequences of drinking spanned individual, interpersonal, and socio-cultural levels. Using alcohol as a means to cope with emotional and financial stress and to facilitate social connection were identified as key motivations. The consequences of fathers’ alcohol use included father health issues, family conflict, and child distress. Barriers to care included limited awareness and lack of access. Importantly, these barriers compounded motivations and consequences of alcohol use. Results can inform both clinical and implementation strategies to better engage and treat fathers’ alcohol use problems. Findings point to the need for a multi-pronged approach to care that considers family dynamics, gender norms, social connections, and individual vulnerabilities and strengths.

Findings suggest multiple motivations for fathers’ drinking. This aligns with the extant literature that engaging in alcohol use can be at times driven by both avoidant coping as well as in response to positive emotion (e.g., celebration, connection). Intervention efforts might consider discussing specific reasons for alcohol use as well as providing steps for adaptive coping, a common element of cognitive behavioral therapies. Interpersonal motivations for alcohol use such as family conflict have implications for family-focused treatments with fathers. First, family-based interventions would benefit from assessing parental mental and behavioral health and well-being in order to provide necessary support. This aligns with the broader developmental literature that suggests that treating caregiver mental health can support positive family outcomes (Cuijpers et al., 2015). Second, to address father alcohol use and related problems, engaging the family system in therapy might facilitate fathers’ treatment engagement (implementation strategy) and support efforts for change. In US based work for instance, couples-based alcohol use interventions have shown positive outcomes for improving both individual alcohol use and couple relationship quality (Lam et al., 2009).

Gender norms around masculinity influenced fathers’ motivation for drinking, consequences of drinking, and barriers to help seeking. Participants reported a pressure among fathers to conform to rigid and hegemonic ideas of masculinity, such as the idea that the only way to be a masculine man is to provide financially. This pressure precipitated distress around not being able to fulfill traditional gender roles and motivated fathers’ alcohol use. Norms then worsened family level conflict, intensified feelings of shame and internalized stigma, and directly interfered with help seeking. The importance of masculinity is consistent with prior work in Eldoret, and globally, that highlights links between ideas around what it means to be a man and patterns of alcohol use and help seeking (Giusto et al., 2022.; Patel et al., 2020). Highlighting the intersection of masculinity and social norms on behavior, for example, a study in Uganda found that men who perceived male peers to drink were more likely to drink themselves (Perkins et al., 2022). At the implementation level, strategies that consider outer-level factors like masculinity are needed. This might include recruitment strategies that consider where men already accept or seek help and delivery approaches that train other fathers to model care engagement as aligned with conceptions of masculinity (Berger et al., 20121203; Giusto et al., 2021). Within treatment itself, exploring the ways in which rigid gender norms influence fathers’ distress and self-conceptions may be critical for addressing symptoms (Giusto et al., 2022; Shafer et al., 2019). Similarly, these approaches might consider the role of poverty and unemployment on fathers’ distress, family-level consequences of alcohol use, and barriers to care. For instance, inclusion of economic empowerment programming combined with treatment may increase engagement and the potential benefit of programming on father outcomes.

The reported consequences of drinking largely centered on interpersonal and family issues. Fathers’ alcohol use had a cascade of effects on the family system, from quarreling, potential for more violent interactions, and youth problems such as missing school. Results align with studies that demonstrate direct and indirect impacts of fathers’ wellbeing on youth mental health (Andreas & O’Farrell, 2007; Schacht et al., 2009). Another reported consequence included the financial impact of fathers’ alcohol use on the family system. Money required for food, school fees, and other basic needs was instead spent on alcohol. This reflects a qualitative study from Eldoret in which participants reported that alcohol use impeded fathers’ financial progress, thereby driving emotional distress and drinking to cope (Patel et al., 2020). Intervention and implementation strategies can support fathers in navigating financial stress to enhance treatment and engage and retain men in care. For instance, a behavioral activation intervention for fathers’ depression and alcohol use in Eldoret incorporated financial tracking (i.e., money saved and spent on alcohol) alongside tracking mood and alcohol use (Giusto et al., 2020). In qualitative interviews following the intervention, fathers commented that seeing how much they saved motivated them to continue reducing their alcohol use. Vocational skills and employment programs could also be important components for engaging men in care, as well as possibly improving mental health and alcohol use outcomes.

Stigma was a key barrier to seeking help and engaging in care. This finding mirrors studies showing stigma as an almost ubiquitous, global barrier to mental health care (Corrigan, 2004; Keyes et al., 2010; Wainberg et al., 2016). Here, mental health and alcohol use stigma intersect social norms around masculinity and identity, compounding its impact. Recommendations for addressing stigma include increasing awareness around mental health and alcohol use problems. Strategies might involve education and sensitization, community advocacy and mobilization, and/or universal advertising of meetings for all people regardless of whether they are impacted by alcohol use (Rao et al., 2019; Thornicroft et al., 2016). A study in Uganda demonstrated that although women are more likely than men to attend community sensitization meetings, they likely pass along meeting information to men through informal networks (Kakuhikire et al., 2021). Other strategies can include working with individuals with lived experience (e.g., fathers in recovery from alcohol use problems) to deliver programs and front national campaigns focused on stigma reduction.

Limitations to this study should be considered when interpreting findings. First, fathers with problem alcohol use in the sample had either previously received or were currently receiving care. This limits our understanding of how fathers who have not received care perceive motivations for alcohol use, the consequences of such use, and barriers to care. Relatedly, the sample did not include partners or children of men experiencing problem alcohol use, which could have provided valuable perspectives.

## Conclusion

This qualitative study highlights the perspectives of multiple community stakeholders on the interrelated factors driving fathers’ alcohol use and the consequences of alcohol use within individual, family, and societal levels. Further, it describes the multiple barriers to care among fathers in Kenya. Fathers often use alcohol to cope with mental health issues, stress, and idleness, and these patterns of use are exacerbated by societal expectations and drinking norms. Consequences of excessive alcohol use include health problems, family conflict, financial strain, and societal stigmatization. Barriers to care include a lack of awareness and trust in available services, accessibility issues, and pervasive stigma. Results can inform multifaceted implementation and intervention approaches to engage fathers in services and effectively treat alcohol use and mental health problems.

## Informed Consent Statement

Informed consent was obtained from all subjects involved in the study.

## Data Availability

Upon reasonable request, deidentified data may be shared.

## Acknowledgments

We acknowledge all the participants who dedicated their time to participate in this study.

